# Non-invasive ^2^H-MRS reveals a greater liver fat contribution from de novo lipogenesis in South Asians compared with Europeans

**DOI:** 10.1101/2025.11.01.25339296

**Authors:** Mueed Azhar, Jieniean Worsley, Kaitlyn M.J.H. Dennis, Emanuella De Lucia Rolfe, Amy Barrett, Mandour O. Mandour, Enes Demir, Katherine Carr, Michele Ferraro, Ranalie De Jesus, Samuel King, Sherly Jose, Simon R. White, Peter Barker, Graham J. Kemp, Kevin M. Brindle, Krishna K. Chatterjee, Nita G. Forouhi, Michelle Venables, Laura Watson, Leanne Hodson, David B. Savage, Alison Sleigh

**Author notes:** Correspondence: Alison Sleigh, Box 65 Addenbrooke’s Hospital, School of Clinical Medicine, Cambridge Biomedical Campus, Cambridge, CB2 0QQ, UK.

## Abstract

Understanding the role of de novo lipogenesis (DNL) in human liver fat accumulation and insulin resistance has been hampered by a lack of non-invasive techniques capable of quantifying DNL-derived liver lipid. Here we develop a precise method that utilises deuterium magnetic resonance imaging, capable of detecting human ^2^H liver lipid signal changes in vivo due to DNL. We formulate MR-specific DNL equations and use these to determine if DNL accounts for the increased liver fat and metabolic risk previously reported in South Asians, compared with age- and BMI- matched individuals from European ancestry. We find an increased fraction of liver fat originates from DNL in South Asians, and that this strongly relates to the amount of liver fat and composition, implying DNL or related factors could play a pivotal role in driving the increased liver fat in South Asians.

## Introduction

South Asians (SA), who constitute a quarter of the world’s population, have a significantly higher risk of developing type 2 diabetes and cardiovascular disease than their European white-origin (E) counterparts with the same body mass index (BMI) (1, 2). Understanding this higher risk is a global health priority. A meta-analysis (2) found that SA accumulate more liver fat than Europeans, even at lower BMI, and as liver fat is a strong predictor of insulin resistance (3–5), this might explain the disparity of metabolic risk between SA and E (2). What remains unclear is the cause of the increased liver fat, which can originate from fatty acids derived from adipose tissue, diet, or synthesis via de novo lipogenesis (DNL).

One hypothesis to explain the higher liver fat in SA, and hence the higher metabolic risk, is the concept of ‘lipid overspill’, whereby hypertrophic subcutaneous adipocytes fail to store fatty acids (FA) properly, resulting in FA being transferred to ‘ectopic’ sites such as liver (6–9). An alternative hypothesis is suggested by the observation (10) that young healthy SA have increased lipogenic sensitivity to dietary sugar, despite normal insulin sensitivity, which indicates DNL may play a role in being an early pathogenic driver of insulin resistance (IR) in SA. In addition to its direct action to increase hepatic triglyceride (TG), elevated rates of DNL can also suppress mitochondrial FA oxidation via its malonyl-CoA intermediate (11), thus also compounding hepatic TG storage. The end product of DNL is usually a saturated FA, and a recent study by Roumans et al. (12) in obesity has shown that increased liver fat is associated with a higher proportion of saturated liver fat, which was in turn associated with hepatic insulin resistance.

Although ^1^H-MRS can non-invasively measure liver fat content and composition (12), it cannot distinguish the component derived from DNL. Liver biopsy is invasive and DNL in humans is traditionally studied using blood-sample tracer studies (13), which offers only indirect estimates of the impact of DNL on liver fat. A more direct non-invasive approach would be useful. Studies performed decades ago in rats showed that ^2^H-MRS could detect turnover of abdominal lipid after cessation of drinking deuterated water (14, 15), however hardware constraints on clinical systems has limited investigation in humans. Recent landmark studies (16, 17) of deuterium metabolic spectroscopy and imaging (DMI) using ^2^H-labelled glucose to study oxidative and glycolytic pathways have sparked an avalanche of interest in deuterium MRS, making suitable hardware now available. Studies measuring hepatic export of DNL-derived palmitate in plasma VLDL or triglyceride typically use ingested deuterated water as the precursor label, aiming at total body water enrichment of ∼0.3 % (18). Over a 5-day labelling period this would theoretically yield DNL-generated changes in ^2^H-liver lipid that are similar to, or less than, the natural abundance ^2^H liver lipid, necessitating the development of ^2^H-DMI techniques that are capable of detecting precise natural abundance liver fat signals.

Here we describe the development of such an MR methodology and derive equations for two MRS liver fat concepts: fractional DNL and absolute DNL, that reflect the fraction of liver fat and the absolute amount of liver fat derived from DNL, respectively. We assess the precision of measuring ^2^H-lipid and compare natural abundance ^2^H-MRS measures of liver fat to standard ^1^H-MRS measures. We show that DNL-derived liver fat is higher in South Asian than in age- and BMI-matched Europeans.

## Results

To ensure time for sufficient fat labelling, and to collect data representative of typical physiological variations, the protocol involved ^2^H water enrichment maintained for 5 days between two visits to the Cambridge Biomedical Campus (see Methods).

### Precise liver ^2^H-lipid measures enables detection of DNL-derived liver lipids

As it is crucial that the ^2^H measure of liver fat is sufficiently precise to support estimates of DNL, the precision was estimated by comparing two halves of the ^2^H fat measurement, termed scan1 and scan2, in all the participants (n=13; 7 SA, 6 E). One SA participant withdrew after Visit 1 such that on Day 5 (n=12; 6 SA, 6 E). A comparison of the two scans showed that ^2^H fat signal repeatability was excellent with an intraclass correlation coefficient (ICC) 0.998 for Day 0 pre-dose (Fig. 1A) and 0.956 for Day 5 post-dose (Fig. 1B). Inter-visit reproducibility (same operator & analyst) over 4 sessions without dosing yielded a CoV of 4.7 % for the ^2^H water signal measurement (data not shown). An example ²H-MRS liver fat spectrum (Fig. 1D) illustrates the post-dose increase in the fat signal. The ^2^H methyl (CH_3_) and methylene (CH_2_) fat peaks are indistinguishable, so the ^2^H fat signal is the integral of the area under both peaks.

**Figure 1.**
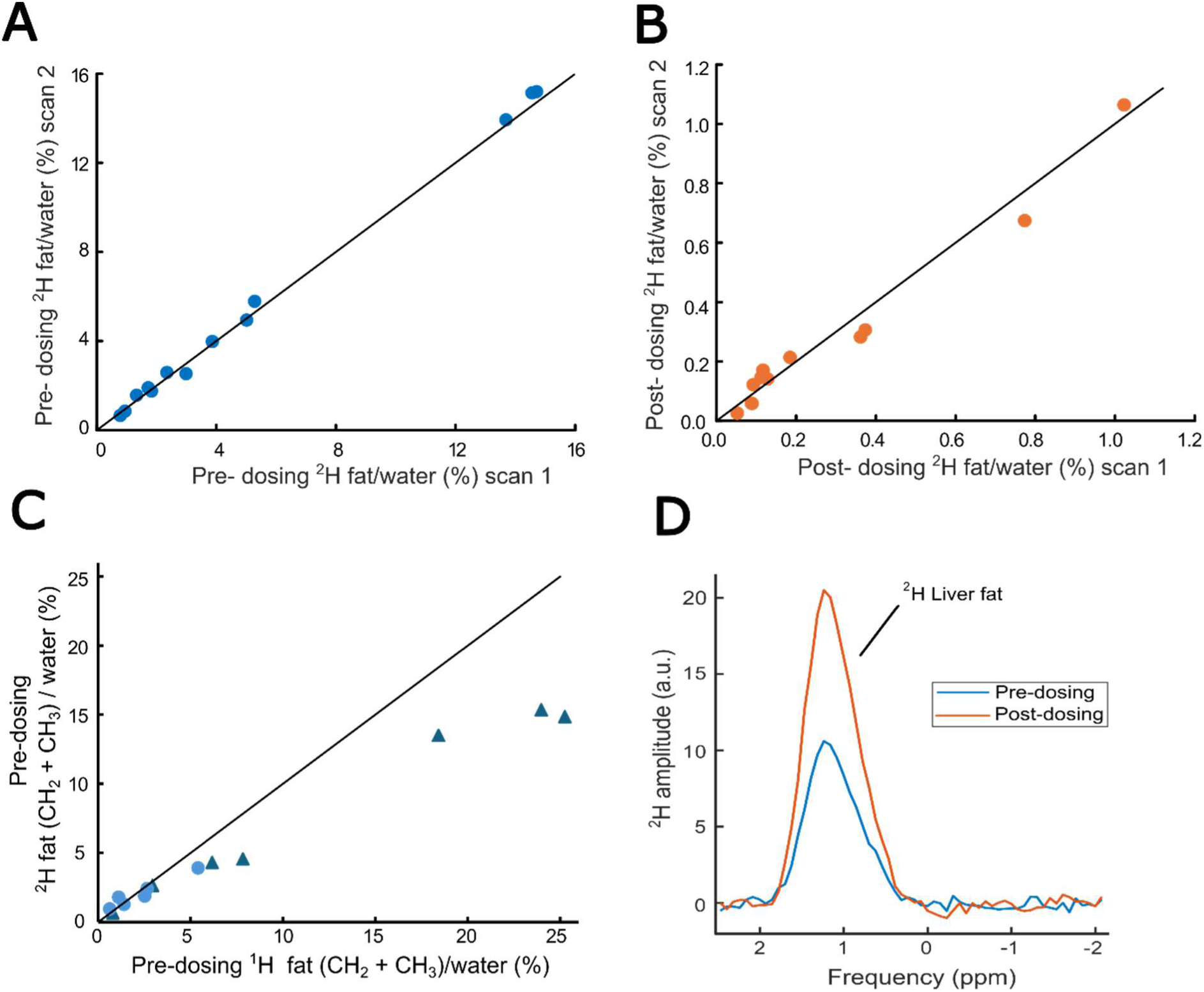
Feasibility of ^2^H-MRS to detect and measure intrahepatic fat. Reproducibility of ²H hepatic fat/water measurements pre-dose (A) and post-dose (B), assessed as comparison of scan 1 and 2 in South Asians and Europeans together. (C) Comparison of fat/water by ^1^H-MRS vs ^2^H-MRS in South Asian (blue triangles) and Europeans (blue circles). (D) Representative ²H-MRS hepatic fat spectra pre-dose (blue) and post-dose (orange), illustrating increase in the fat signal post-dose. In (A-C) the solid line represents identity.

The accuracy of the ^2^H fat signal measurement was investigated by comparing the pre-dose (natural abundance) ^2^H-MRS fat/water signal ratio with the equivalent ^1^H-MRS measure (CH_2_+CH_3_)/water in SA and E (Fig. 1C). The measures were highly correlated (r_s_ = 0.984, p = 0.001; SA and E combined), however, interestingly the points deviated from the line of identity at higher liver fat, demonstrating a lower natural abundance of ^2^H fat/water ratio compared with ^1^H. This ^2^H/^1^H ratio of fat/water was lower in SA *vs* E (p = 0.022), with median 0.70 *vs* 0.91.

### Liver fat is higher in South Asians than in Europeans

The SA and E participants were age- and BMI-matched (Table 1) and spanned a large BMI range: SA (23.6 – 40.8 kg/m^2^) and E (24.9 – 43.2 kg/m^2^); none had diabetes. By ^1^H MRS, intrahepatic lipid (IHL) content (p = 0.035), but not saturation, was higher in SA *vs* E.

**Table 1.**
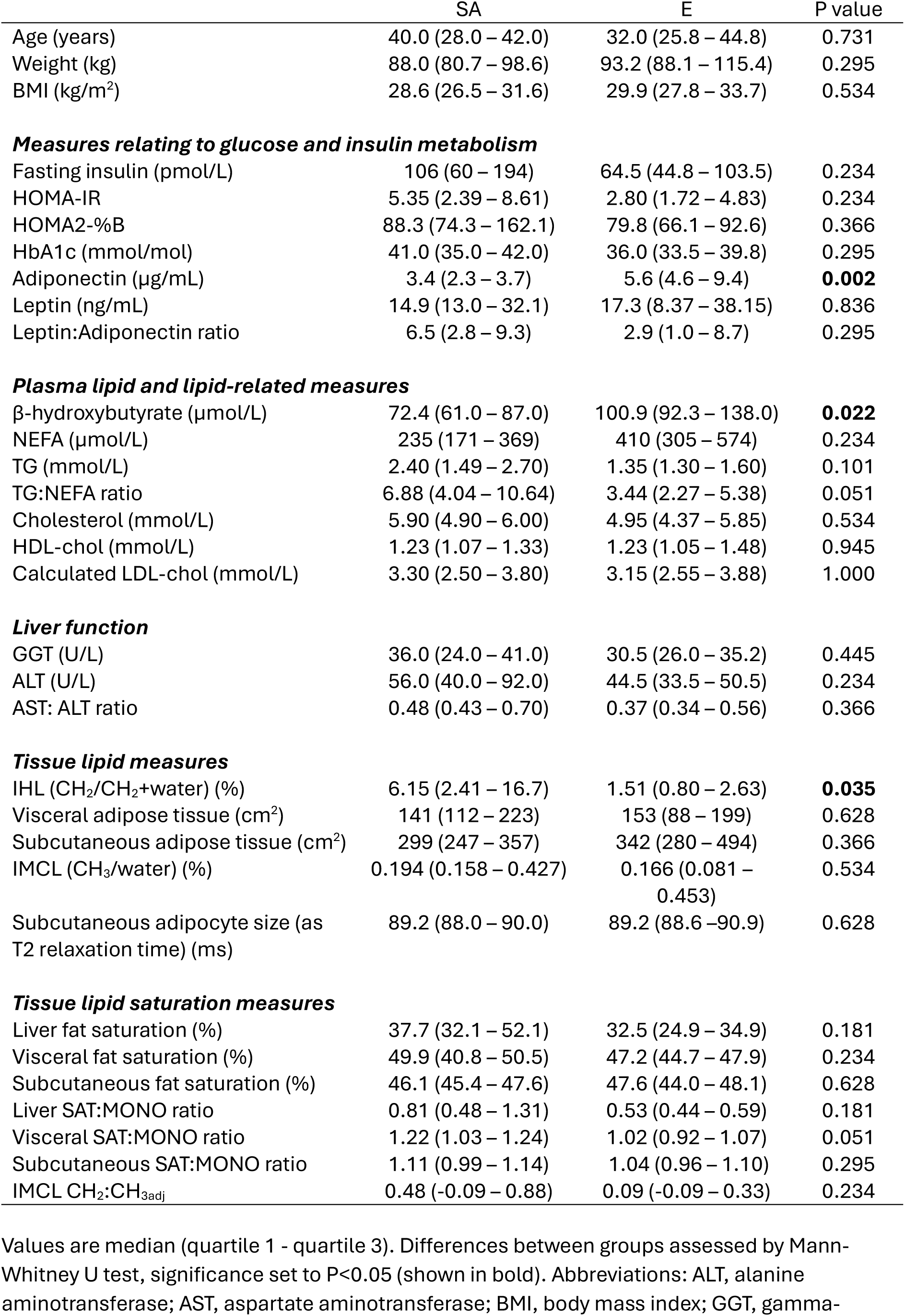
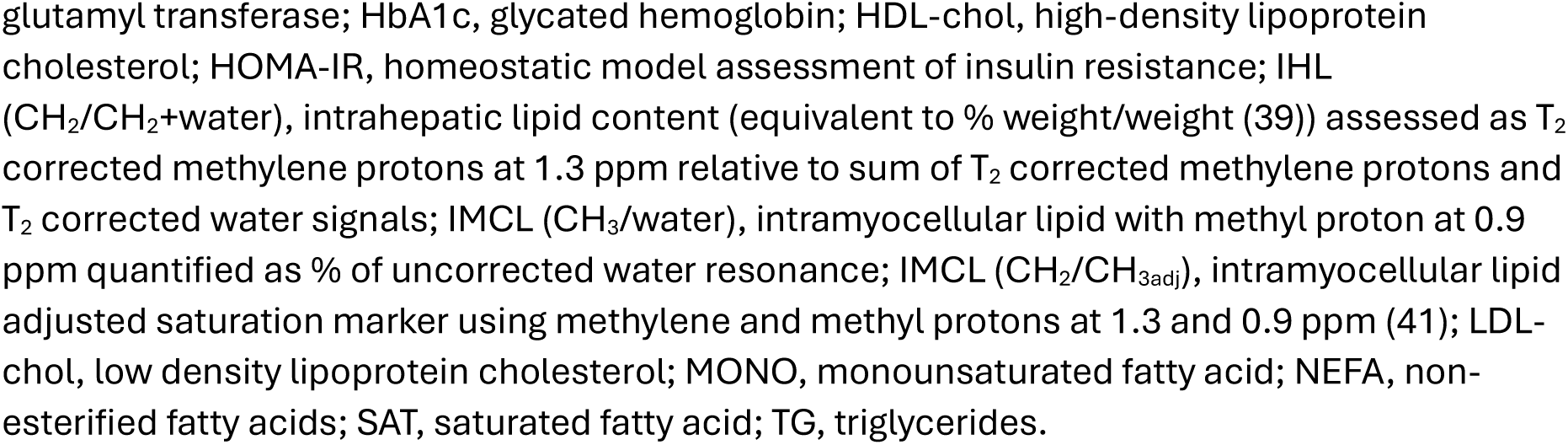
Participant characteristics in South Asians (SA) and Europeans (E)

|  | SA | E | P value |
| --- | --- | --- | --- |
| Age (years) | 40.0 (28.0 – 42.0) | 32.0 (25.8 – 44.8) | 0.731 |
| Weight (kg) | 88.0 (80.7 – 98.6) | 93.2 (88.1 – 115.4) | 0.295 |
| BMI (kg/m<sup>2</sup>) | 28.6 (26.5 – 31.6) | 29.9 (27.8 – 33.7) | 0.534 |
| ***Measures relating to glucose and insulin metabolism*** |  |  |  |
| Fasting insulin (pmol/L) | 106 (60 – 194) | 64.5 (44.8 – 103.5) | 0.234 |
| HOMA-IR | 5.35 (2.39 – 8.61) | 2.80 (1.72 – 4.83) | 0.234 |
| HOMA2-%B | 88.3 (74.3 – 162.1) | 79.8 (66.1 – 92.6) | 0.366 |
| HbA1c (mmol/mol) | 41.0 (35.0 – 42.0) | 36.0 (33.5 – 39.8) | 0.295 |
| Adiponectin (µg/mL) | 3.4 (2.3 – 3.7) | 5.6 (4.6 – 9.4) | **0.002** |
| Leptin (ng/mL) | 14.9 (13.0 – 32.1) | 17.3 (8.37 – 38.15) | 0.836 |
| Leptin:Adiponectin ratio | 6.5 (2.8 – 9.3) | 2.9 (1.0 – 8.7) | 0.295 |
| ***Plasma lipid and lipid-related measures*** |  |  |  |
| β-hydroxybutyrate (µmol/L) | 72.4 (61.0 – 87.0) | 100.9 (92.3 – 138.0) | **0.022** |
| NEFA (µmol/L) | 235 (171 – 369) | 410 (305 – 574) | 0.234 |
| TG (mmol/L) | 2.40 (1.49 – 2.70) | 1.35 (1.30 – 1.60) | 0.101 |
| TG:NEFA ratio | 6.88 (4.04 – 10.64) | 3.44 (2.27 – 5.38) | 0.051 |
| Cholesterol (mmol/L) | 5.90 (4.90 – 6.00) | 4.95 (4.37 – 5.85) | 0.534 |
| HDL-chol (mmol/L) | 1.23 (1.07 – 1.33) | 1.23 (1.05 – 1.48) | 0.945 |
| Calculated LDL-chol (mmol/L) | 3.30 (2.50 – 3.80) | 3.15 (2.55 – 3.88) | 1.000 |
| ***Liver function*** |  |  |  |
| GGT (U/L) | 36.0 (24.0 – 41.0) | 30.5 (26.0 – 35.2) | 0.445 |
| ALT (U/L) | 56.0 (40.0 – 92.0) | 44.5 (33.5 – 50.5) | 0.234 |
| AST: ALT ratio | 0.48 (0.43 – 0.70) | 0.37 (0.34 – 0.56) | 0.366 |
| ***Tissue lipid measures*** |  |  |  |
| IHL (CH<sub>2</sub>/CH<sub>2</sub>+water) (%) | 6.15 (2.41 – 16.7) | 1.51 (0.80 – 2.63) | **0.035** |
| Visceral adipose tissue (cm<sup>2</sup>) | 141 (112 – 223) | 153 (88 – 199) | 0.628 |
| Subcutaneous adipose tissue (cm<sup>2</sup>) | 299 (247 – 357) | 342 (280 – 494) | 0.366 |
| IMCL (CH<sub>3</sub>/water) (%) | 0.194 (0.158 – 0.427) | 0.166 (0.081 – 0.453) | 0.534 |
| Subcutaneous adipocyte size (as T2 relaxation time) (ms) | 89.2 (88.0 – 90.0) | 89.2 (88.6 – 90.9) | 0.628 |
| ***Tissue lipid saturation measures*** |  |  |  |
| Liver fat saturation (%) | 37.7 (32.1 – 52.1) | 32.5 (24.9 – 34.9) | 0.181 |
| Visceral fat saturation (%) | 49.9 (40.8 – 50.5) | 47.2 (44.7 – 47.9) | 0.234 |
| Subcutaneous fat saturation (%) | 46.1 (45.4 – 47.6) | 47.6 (44.0 – 48.1) | 0.628 |
| Liver SAT:MONO ratio | 0.81 (0.48 – 1.31) | 0.53 (0.44 – 0.59) | 0.181 |
| Visceral SAT:MONO ratio | 1.22 (1.03 – 1.24) | 1.02 (0.92 – 1.07) | 0.051 |
| Subcutaneous SAT:MONO ratio | 1.11 (0.99 – 1.14) | 1.04 (0.96 – 1.10) | 0.295 |
| IMCL CH<sub>2</sub>:CH<sub>3adj</sub> | 0.48 (-0.09 – 0.88) | 0.09 (-0.09 – 0.33) | 0.234 |

Values are median (quartile 1 - quartile 3). Differences between groups assessed by Mann-Whitney U test, significance set to P<0.05 (shown in bold). Abbreviations: ALT, alanine aminotransferase; AST, aspartate aminotransferase; BMI, body mass index; GGT, gamma-
glutamyl transferase; HbA1c, glycated hemoglobin; HDL-chol, high-density lipoprotein cholesterol; HOMA-IR, homeostatic model assessment of insulin resistance; IHL ( $\text{CH}_2/\text{CH}_2+\text{water}$ ), intrahepatic lipid content (equivalent to % weight/weight (39)) assessed as $T_2$ corrected methylene protons at 1.3 ppm relative to sum of $T_2$ corrected methylene protons and $T_2$ corrected water signals; IMCL ( $\text{CH}_3/\text{water}$ ), intramyocellular lipid with methyl proton at 0.9 ppm quantified as % of uncorrected water resonance; IMCL ( $\text{CH}_2/\text{CH}_{3\text{adj}}$ ), intramyocellular lipid adjusted saturation marker using methylene and methyl protons at 1.3 and 0.9 ppm (41); LDL-chol, low density lipoprotein cholesterol; MONO, monounsaturated fatty acid; NEFA, non-esterified fatty acids; SAT, saturated fatty acid; TG, triglycerides.

There were no differences in abdominal visceral or subcutaneous adipose tissue volumes (both p>0.4) nor subcutaneous adipocyte size as assessed indirectly by the methylene transverse (T_2_) relaxation time (p = 0.6). The T_2_ relaxation time relates to the magnetic resonance signal decay over time and is significantly influenced by the local environment, particularly molecular motion. The methylene T_2_ relaxation time has been shown to correlate with median lipid droplet diameter irrespective of lipid composition (19). Circulating β-hydroxybutyrate (BHB) and adiponectin were lower in SA *vs* E (p = 0.022 and p = 0.002 respectively).

### DNL-derived liver lipid is higher in South Asians than Europeans, and relates to liver fat content and composition

The fractional and absolute DNL_MR_ were higher in SA *vs* E (Fig. 2A & B; both p = 0.041). They showed significant positive correlations with IHL content (fractional DNL_MR_ r_s_ = 0.874, p = 0.001, Fig 2C; absolute DNL_MR_ r_s_ = 0.888, p = 0.001, Fig 2D), as well as with the SAT:MONO ratio of both IHL and VAT (Table 2). They also both showed a significant positive correlation with gamma-glutamyl transferase, GGT (fractional DNL_MR_ r_s_ = 0.632, p = 0.028; absolute DNL_MR_ r_s_ = 0.600, p = 0.039) and a negative correlation with adiponectin (fractional DNL_MR_ r_s_ = -0.695, p = 0.012; absolute DNL_MR_ r_s_ = - 0.705, p = 0.010); all n = 12. A negative correlation with BHB (fractional: r_s_ = -0.506, p = 0.093; absolute: r_s_ = -0.541, p = 0.069; Figs 2E & F) fell short of statistical significance.

**Figure 2.**
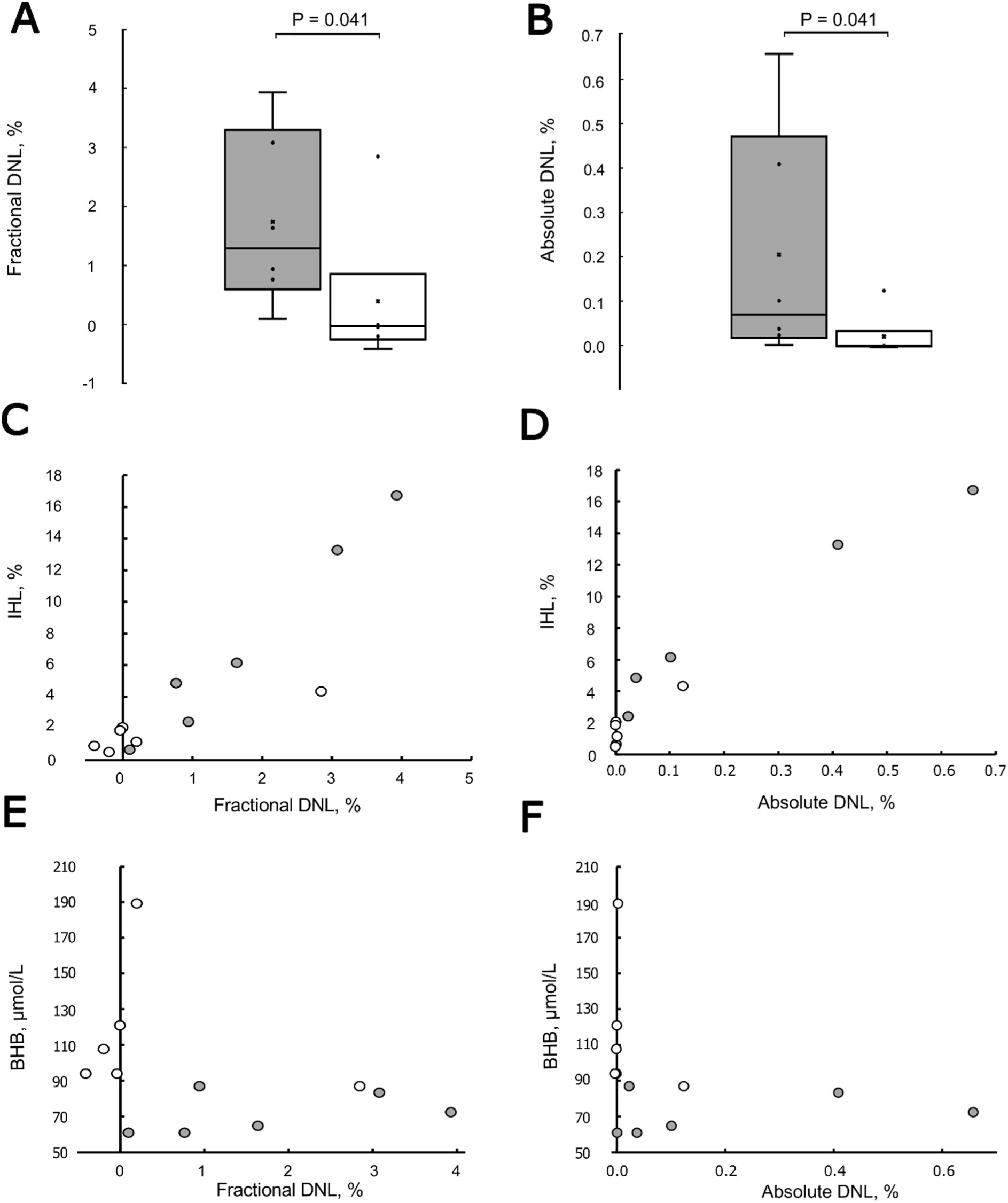
De novo lipogenesis (DNL) derived liver fat and intrahepatic lipid (IHL) measures in South Asian (SA) and Europeans (E). Box and whisker plot for (A) fractional and (B) absolute DNL measures in SA (grey bars) and E (white bars). (C-F) Comparison of fractional DNL with IHL (C) and circulating β-hydroxybutyrate (E); comparison of absolute DNL with IHL (D) and β-hydroxybutyrate BHB (F), in SA (grey circles) and E (white circles).

**Table 2.** Associations of liver content and MR DNL metrics with lipid saturation indices.

|  | [IHL] | Fractional DNL | Absolute DNL | <sup>2</sup>H/<sup>1</sup>H ratio fat/water |
| --- | --- | --- | --- | --- |
| [IHL] | - | 0.874 *** | 0.888 *** | - 0.861 *** |
| IHL SAT:MONO ratio | 0.775 ** | 0.692 * | 0.748 ** | - 0.792 *** |
| VAT SAT:MONO ratio | 0.792 *** | 0.648 * | 0.708 ** | - 0.814 *** |
| IMCL CH<sub>2</sub>:CH<sub>3adj</sub> ratio | 0.225 | 0.266 | 0.336 | - 0.600 * |
| SCAT SAT:MONO ratio | 0.154 | 0.413 | 0.441 | - 0.371 |

The table shows Spearman's correlation coefficients. \* $p \leq 0.05$ ; \*\* $p \leq 0.01$ ; \*\*\* $p \leq 0.001$
Abbreviations; DNL, de novo lipogenesis; [IHL], intrahepatic lipid content assessed as (CH<sub>2</sub>/[CH<sub>2</sub>+water]); IMCL (CH<sub>2</sub>/CH<sub>3adj</sub>), intramyocellular lipid adjusted saturation marker using methylene and methyl protons at 1.3 and 0.9 ppm (41); MONO, monounsaturated fatty acid; SAT, saturated fatty acid; SCAT, subcutaneous adipose tissue; VAT, visceral adipose tissue.

The pre-dose ^2^H/^1^H ratio of fat/water strongly correlated negatively with both fractional DNL_MR_ (r_s_ = -0.809, p = 0.001, n = 12) and absolute DNL_MR_ (r_s_ = -0.844, p = 0.001, n = 12). Similar to DNL_MR_ measures, but inversely, pre-dose ^2^H/^1^H ratio of fat/water correlated negatively with IHL content (r_s_ = -0.861, p = 0.001, Table 2), liver and VAT SAT:MONO ratio (Table 2), GGT (r_s_ = 0.666, p = 0.013) and adiponectin (r_s_ = - 0.600, p = 0.030); all n = 13. Pre-dose ^2^H/^1^H ratio of fat/water also correlated negatively with IHL saturation (r_s_ = -0.638, p = 0.019), VAT volume (r_s_ = -0.716, p = 0.006), and the intramyocellular lipid (IMCL) compositional saturation index (CH_2_:CH_3adj_) (r_s_ = -0.600, p = 0.030; Table 2).

IHL and DNL_MR_ measures did not significantly correlate with measures of non-hepatic lipid pool size, either anatomical (SCAT, VAT) or intra-organ (IMCL). IHL content significantly correlated with IHL SAT:MONO and VAT SAT:MONO, but not SCAT SAT:MONO (p = 0.6) (Table 2).

### Labelling effectiveness and compliance

Plasma ^2^H water enrichment above baseline was 0.292 ± 0.015 % (mean ± SEM; SA & E combined) on Day 1 (12h post initial priming dose), close to our intended target enrichment of 0.3 %. Maintenance water diaries (see methods) and returned empty ^2^H_2_O vials indicated that participants were compliant, with an average of 2.3 ± 0.1 litres of maintenance water consumed per day, and only 6.3 ± 1.8 % of drinking liquid volume reported from non-maintenance water. Despite maintenance water being enriched slightly higher (0.45 %), average plasma ^2^H water enrichment above baseline on Day 5 was slightly lower at 0.263 ± 0.013 (p = 0.01; paired t-test) than on Day1.

### Relationships to plasma DNL (DNL_plasma_) measures

Taking all participants together, Day 5 DNL_plasma_ showed a significant positive correlation with MR-measured fractional DNL (r_s_ = 0.664, p = 0.018; Supplementary S1B), absolute DNL (r_s_ = 0.650, p = 0.022), and a negative correlation with ^2^H/^1^H ratio of fat/water (r_s_ = -0.581, p = 0.047). Day 1 DNL_plasma_ did not significantly correlate with MR-measured fractional or absolute DNL (both p > 0.3; supplementary S1A). Day 5, but not Day 1, DNL_plasma_ correlated positively with liver SAT:MONO (r_s_ = 0.643, p = 0.024), and a positive correlation with IHL content fell short of statistical significance (r_s_ = 0.573, p = 0.051; Supplementary S1C). Day 1 and Day 5 DNL_plasma_ showed no significant associations with any biochemical measure other than positive correlations with leptin on Day 1 (r_s_ = 0.681, p = 0.01) and GGT on Day 5 (r_s_ = 0.761, p = 0.004).

### Relationship of liver fat to indirect measures assessing the ‘lipid overspill’ hypothesis

We did not directly assess lipid spillover, however, fasting NEFA concentration as an indirect measure of adipose tissue lipolysis, was not significantly related to liver fat (p = 0.5), nor higher in SA *vs* E (Table 1). Liver fat was not significantly related to subcutaneous adipocyte size, measured indirectly as the methylene T_2_ relaxation time (19) (p = 0.2), and this did not differ in SA *vs* E (p = 0.6). Subcutaneous adipocyte size correlated positively with SCAT volume (r_s_ = 0.610, p = 0.027; Figure 3A), but SA did not have significantly higher adipocyte size for a given SCAT volume (Figure 3A).

**Figure 3.**
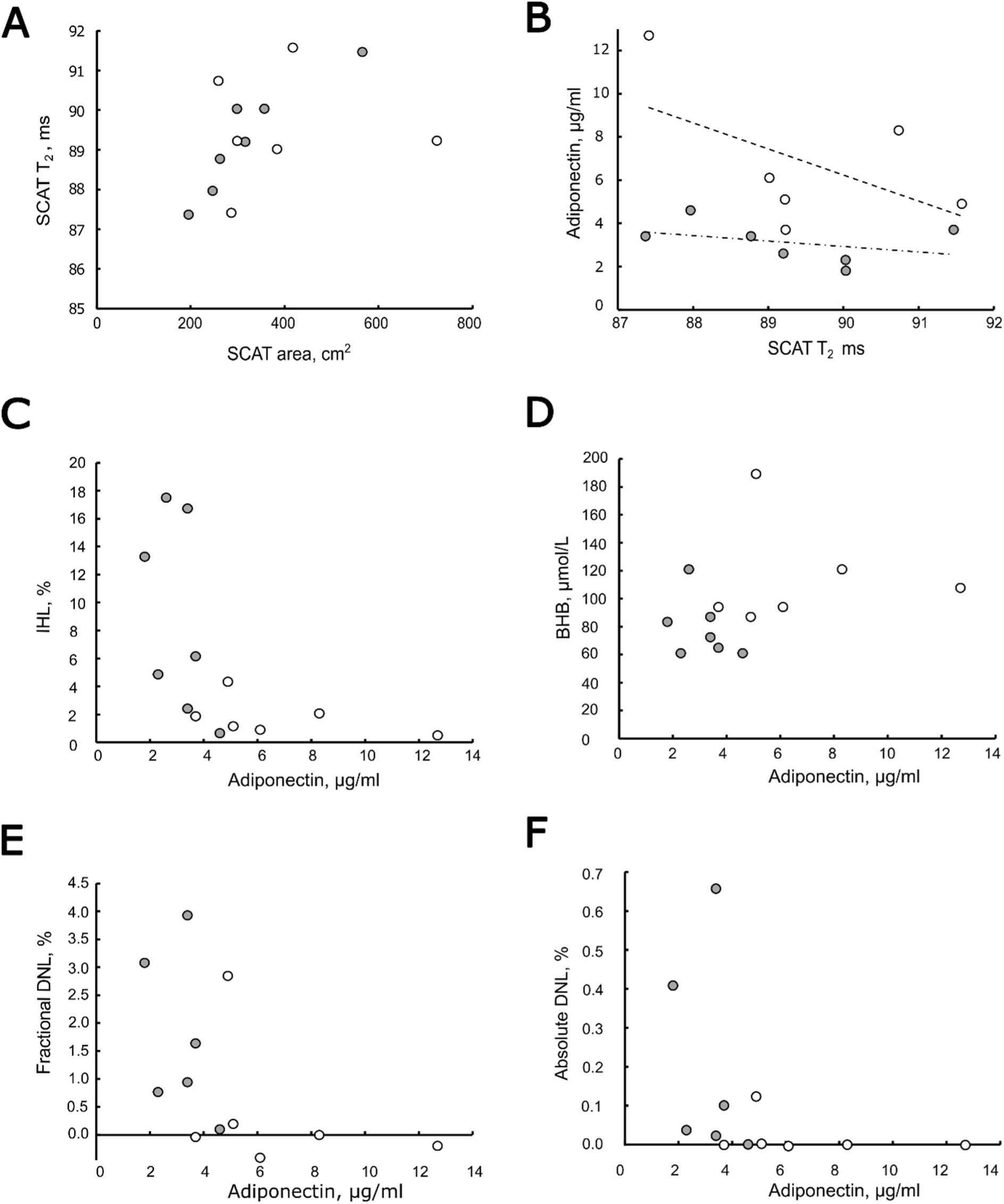
Subcutaneous adipose tissue measures and associations of adiponectin levels with metabolic markers related to hepatic lipid metabolism in South Asian (SA) and Europeans (E). Relationships of subcutaneous adipocyte size, assessed by the T2 relaxation time, to subcutaneous adipose tissue volume (A) and adiponectin (B). Relationships of adiponectin to intrahepatic lipid (C), β-hydroxybutyrate BHB (D), fractional DNL (E), and absolute DNL (F). SA are grey circles and E white circles. Dashed and dash-dot lines in (B) are regression lines through the SA and E datapoints, respectively. Abbreviations: [IHL], intrahepatic lipid content assessed as (CH_2_/CH_2_+water); SCAT, subcutaneous adipose tissue.

### Relationships with adiponectin

Taking all participants together, circulating adiponectin showed a significant negative correlation with IHL (r_s_ = -0.771, p = 0.002; Figure 3C) and DNL_MR_ (fractional: r_s_ = -0.695, p = 0.012; absolute: r_s_ = -0.705, p = 0.010; Figures 3E & F), and correlated positively with pre-dose MR ^2^H/^1^H ratio of fat/water (r_s_ = 0.600, p = 0.030). Adiponectin correlated negatively with TG (r_s_ = -0.576, p = 0.040) and glucose (r_s_ = -0.653, p = 0.015). A positive correlation with BHB fell short of statistical significance (r_s_ = 0.496, p = 0.085; Figure 3D). Figure 3B shows the relationship of SCAT adipocyte size and adiponectin in SA and E, and for a given adipocyte size SA had consistently lower adiponectin. Similarly, for a given SCAT volume, SA had consistently lower adiponectin.

### HOMA-IR relates to liver fat content and DNL

Taking all participants together, HOMA-IR showed a significant positive correlation with IHL content (r_s_ = 0.659, p = 0.014, n = 13) but no relation with IHL saturation or IHL SAT:MONO ratio (both p> 0.4). HOMA-IR had a positive correlation with both fractional (r_s_ = 0.552, p = 0.06, n = 12) and absolute DNL (r_s_ = 0.524, p = 0.08, n = 12) that fell short of statistical significance, and a negative correlation with pre-dose ^2^H/^1^H ratio of fat/water (r_s_ = -0.547, p = 0.05, n = 13). Of the biochemical measurements, HOMA-IR showed a significant positive correlation with TG (r_s_ = 0.559, p = 0.047), GGT (r_s_ = 0.793, p = 0.001), and ALT (r_s_ = 0.676, p = 0.011), and a negative correlation with adiponectin fell short of statistical significance (r_s_ = - 0.504, p = 0.079), all n = 13.

### South Asian weight loss case study reveals reduced DNL and liver fat concentration

Following feedback on their liver fat content, one SA participant independently chose to lose weight by calorie restriction; they lost 10 kg over 4 months (BMI 26.5 to 23.5 kg/m^2^), after which they were weight-stable for 2 months before they requested a further set of measurements. Results are shown in Table 3. As expected, there were decreases in VAT and SCAT volume as well as SCAT adipocyte size. Fractional and absolute DNL were dramatically reduced 26 and 93-fold respectively, and liver fat content and saturation concurrently reduced. HOMA-IR and TG decreased, BHB increased, and adiponectin levels slightly increased yet were still low following weight loss.

**Table 3.** Case study of one South Asian (SA) participant before and after weight loss.

|  | Pre-diet | Post-diet | Reference range |
| --- | --- | --- | --- |
| Age (years) | 41.3 | 41.8 | - |
| Weight (kg) | 92.8 | 82.8 | - |
| BMI (kg/m<sup>2</sup>) | 26.5 | 23.7 | 18.5–24.9 |
| HOMA-IR | 1.98 | 1.09 | < 1.0 good (>1.9 early IR) |
| Adiponectin (µg/mL) | 2.3 | 3.6 | 5–30 |
| β-hydroxybutyrate (µmol/L) | 61 | 111 | < 600 |
| NEFA (µmol/L) | 218 | 321 | 300 - 600 |
| TG (mmol/L) | 1.5 | 1.0 | < 1.7 |
| **Lipid measures** |  |  |  |
| IHL (CH<sub>2</sub>/ CH<sub>2</sub> + water) (%) | 4.86 | 1.29 | < 5.5 |
| Visceral adipose tissue (cm<sup>2</sup>) | 87.9 | 77.7 | 12.0 – 92.4 <sup>a</sup> |
| Subcutaneous adipose tissue (cm<sup>2</sup>) | 299 | 209 | 35 – 234 <sup>a</sup> |
| IMCL (CH<sub>3</sub>/water) (%) | 0.57 | 0.27 | 0.03 – 0.47 <sup>a</sup> |
| Subcutaneous adipocyte size (T2 relaxation time) (ms) | 90.0 | 87.2 | - |
| Fractional DNL (of total liver fat) (%) | 0.765 | 0.029 | - |
| Absolute DNL fat/water (%) | 0.0372 | 0.0004 | - |
| **Lipid saturation measures** |  |  |  |
| Saturated liver fat (%) | 52.08 | 26.74 | - |
| Visceral fat saturation (%) | 50.48 | 46.10 | - |
| Subcutaneous fat saturation (%) | 46.80 | 47.37 | - |
| Liver SAT:MONO ratio | 1.31 | 0.51 | - |
| Visceral SAT:MONO ratio | 1.22 | 1.03 | - |
| Subcutaneous SAT:MONO ratio | 1.07 | 1.05 | - |
| IMCL CH<sub>2</sub>:CH<sub>3adj</sub> | 1.55 | 0.40 | -0.21 – 0.56 <sup>a</sup> |

<sup>a</sup> Taken from subset of lean European-white males with BMI<25 kg/m<sup>2</sup> (n=15) from Azhar *et al* (3). Age (mean ± SEM) = 32.4±3.4 years and BMI = 23.1±0.4 kg/m<sup>2</sup>.
Abbreviations: BMI, body mass index; DNL, de novo lipogenesis; HOMA-IR, homeostatic model assessment of insulin resistance; IHL(CH<sub>2</sub>/CH<sub>2</sub>+water), intrahepatic lipid content (equivalent to % weight/weight (39)) assessed using T<sub>2</sub>-corrected methylene protons at 1.3 ppm relative to the sum of T<sub>2</sub> corrected methylene protons and T<sub>2</sub> corrected water signals; IMCL (CH<sub>3</sub>/water), intramyocellular lipid with methyl proton at 0.9 ppm quantified as % of uncorrected water; IMCL (CH<sub>2</sub>/CH<sub>3adj</sub>), intramyocellular lipid adjusted saturation marker using methylene and methyl protons at 1.3 and 0.9 ppm (41); MONO, monounsaturated fatty acid; NEFA, non-esterified fatty acids; SAT, saturated fatty acid; TG, triglycerides.

## Discussion

Hepatic fat is a strong predictor of metabolic disease risk, but understanding the role of DNL in human hepatic fat accumulation and insulin resistance has been hampered by a lack of non-invasive methods capable of quantifying DNL-derived liver lipid. We have developed a method using deuterium magnetic resonance imaging which has excellent precision and is able to detect human ^2^H liver lipid signal changes in vivo due to DNL following 5 days of drinking deuterated water. As expected, the resulting lipid signal changes were small compared with the intersubject range of liver fat (the biggest individual change roughly doubled the natural abundance lipid signal), so consideration of baseline ^2^H lipid signal is essential in evaluating the stored lipid formed from DNL. We give MR-specific equations that do this, accounting for the maximum number of deuterium atoms incorporated into palmitate. We used this to quantify the fraction of liver lipid and the absolute amount of liver lipid arising from DNL over the 5 days of labelling.

Using this method, we have shown that both the fractional and absolute amount of liver lipid from DNL are higher in South Asians (SA) compared with age- and BMI-matched males from European-white ancestry (E). These DNL measures had a striking association with the amount of liver fat, supporting the hypothesis that DNL plays an important role in driving the higher liver fat in SA. Nevertheless, the actual fraction of liver lipid generated by DNL over the 5 days was small. This is not unexpected. While 5 days labelling is enough to generate a quantifiable signal, and is, we argue, a physiologically representative timeframe with respect to diurnal and fasted/fed cycles, it is not long enough to reach equilibrium, as an estimation of the turnover of liver fat is > 1 month (20). The half-life of the liver palmitate pool can vary, but estimations using the decay kinetics for the slow turnover pool of palmitate released from liver into blood yielded an average of 25 days in 103 people with NASH without cirrhosis (21). Very roughly, therefore, multiplying our fractional DNL by 1/(1-e^(-5/25)^) ≈ 5.5 gives an estimate of *actual* fractional DNL – the largest value in our study being 22 % in a participant with IHL 16 % (equivalent to g/g tissue). This is in reasonably consistent with the report (20) that DNL contributed 26±7 % to liver fat in a cohort of NAFLD which had, on average, a liver fat of 16 % (g/g tissue).

Although a 22 % contribution is not negligible, it is clearly not the largest contributor to the liver fat pool, suggesting that the strong association of DNL with liver fat is therefore likely influenced by other interconnected factors. An obvious candidate is malonyl-CoA, an intermediate in DNL that tends to decrease FA oxidation by inhibiting the mitochondrial FA transporter CPT-1. There was a non-significant tendency for β-hydroxybutyric acid (BHB, a surrogate marker of hepatic FA oxidation) to correlate with DNL measures, however, BHB was low in all SA compared to E, irrespective of DNL. Adiponectin levels were significantly lower in SA vs E, consistent with previous reports (1, 22, 23). It has been suggested by others that adiponectin can increase FA oxidation and simultaneously inhibit DNL (24, 25), and if this is so, then this could have significant influence on the relationship of DNL with liver fat. Indeed, adiponectin levels were closely associated with liver fat and liver DNL measures across all participants, with a tendency falling short of statistical significance to correlate also with BHB, the surrogate marker of FA oxidation.

We did not include direct measurement of lipolysis, so could not directly test the hypothesis of ‘lipid overspill’ due to hypertrophic adipocytes. However, the finding that the liver fat fractional DNL over the 5 days is strongly associated with liver fat concentration (i.e. the higher IHL, the larger the proportion originating from DNL) is difficult to reconcile with ‘overspill’ as the main cause of higher liver fat in SA. Consistent with this we found no SA *vs* E differences in circulating FA or subcutaneous adipocyte size or numbers, with no obvious difference in the association of adipocyte size and subcutaneous adipose tissue volume.

Other support for our method comes from the close correlation between the ^2^H natural abundance liver fat to water ratio and the equivalent ^1^H measure. Interestingly, deviation from the line of identity in a graph comparing these measures reflects the natural abundance hydrogen isotopic composition (^2^H/^1^H ratio) of fat relative to water. This can be affected by the H isotope composition of cellular water, acetate, and NADPH (the substrates for FA synthesis), by H isotope fractionation by the enzymes of FA synthesis (which in general prefer lighter isotopologues), as well as isotope fractionation during fatty acid transport and metabolism. As a result, FA formed by DNL are generally ^2^H depleted compared with dietary lipids (26).

The ^2^H/^1^H ratio of fat relative to water was significantly lower in SA *vs* E (median 0.7 *vs* 0.9), which are physiologically plausible given ^2^H fat/water ratios of e.g. 0.86 in olive oil and 0.80 in beef fat (27) and the maximum enrichment of deuterium into newly synthesized palmitate is ∼65 % (28), equating to a maximum ^2^H/^1^H ratio of fat relative to water of 0.65. The ^2^H/^1^H ratio marker was highly correlated with liver fat DNL measures and had similar and stronger associations with liver fat content and adiponectin levels to DNL measures. We speculate that this natural abundance ^2^H/^1^H measure could be an alternative marker of DNL that has the advantage of not requiring dosing, not being dependent on normalisation to an external phantom, and is representative of a large timescale (being equivalent to labelling the whole liver fat pool).

Comparing our MR DNL measures to traditional plasma DNL measures, we found associations with plasma DNL on Day 5 but not Day 1, suggesting that a longer labelling period may be necessary to be representative of the fraction of liver fat originating from DNL. We sample the total plasma triglyceride pool that includes lipoproteins with a range of half-lives (29) that can, together, capture and store labelling information on a timescale that covers both fed and fasted states and more closely matches the MR DNL measurement. The longer labelling period could also be detecting DNL-generated lipids passing through the liver lipid pool and a steady-state recirculation of lipoproteins back to the liver.

The liver fat content, MR DNL and ^2^H/^1^H fat/water measures, and the lipid composition (SAT:MONO) of liver and visceral adipose tissue were all strongly correlated, yet in this study of limited numbers it was the liver fat content and MR DNL rather than liver fat composition that were related to insulin sensitivity assessed by HOMA-IR. Weight loss has previously been shown to reduce DNL assessed by plasma TG after labelling for 3-5 weeks (30). In the SA individual who went on to deliberately lose weight we found striking decreases in liver fat fractional and absolute DNL, as well as an almost near doubling of BHB, the indirect marker of fatty acid oxidation, with a concomitant reduction in liver fat content and saturation, and improvements in insulin sensitivity. Adiponectin increased, but despite BMI now being 23.7 kg/m^2^, it was still lower than all E participants (whose BMI went up to 43.2 kg/m^2^). This is reflected in the whole study where SA had consistently lower adiponectin for a given subcutaneous adipose tissue volume or adipocyte size, than Europeans. It would appear that although lifestyle alterations are able to reverse metabolic sequela, SA individuals with low adiponectin may still be more at risk of metabolic disease in situations of calorie excess. The higher metabolic risk in SA is internationally recognised, with the World Health Organization guidelines recommending a lower BMI threshold of 23 kg/m^2^ for classification of being overweight. As ∼25 % of world’s population are of South Asian origin, reversal of this risk would have significant world-wide health and socioeconomic impact.

Adiponectin is consistently reported as being lower in South Asians (1, 22, 23), and a study in healthy newborns of South Asians aged 3-6 months (31) who had lower adiponectin with normal insulin and lipid profiles concluded that factors affecting lowered adiponectin in early life precede metabolic alterations in later life. Supporting this, lowered adiponectin has been shown to be an independent predictor of T2D in Asian Indians (32), and whilst our study is of limited numbers, it may provide some mechanistic insight of the higher metabolic risk in South Asians. Future studies would be needed to confirm our findings in a larger and more diverse population, as well as in females.

In summary, we have developed a non-invasive method for assessing hepatic lipids made by de novo lipogenesis that utilises precise ^2^H measurements of natural abundance signals and subsequent changes following ingestion of deuterated water to enrichments typically used in studies assessing ^2^H incorporation into plasma palmitate. In this study, which predominantly was in overweight individuals, the proportion of liver fat that originated from DNL was closely related to liver fat content. Together with biochemical information, this has provided mechanistic insight into the higher metabolic risk in South Asians and revealed potential avenues to mitigate the disproportionate risk. Our findings highlight the value of ²H-MRS as a powerful non-invasive tool for investigating human fat metabolism; employing the methodology we describe will enable determination of the role of de novo lipogenesis in a range of metabolic disorders including fatty liver disease.

## Methods

### Participants

We recruited 13 healthy male adult volunteers (7 South Asian and 6 European) matched for age and body mass index (BMI). All participants had lived within a 30-mile radius of the Cambridge Biomedical Campus for at least 1 year prior to study; South Asian participants self-reported to be either South Asian or Indian, Europeans all self-reported to be White British or European. None of the participants were vegetarian or vegan. Exclusion criteria were MRI contraindications, medication known to alter DNL, weekly consumption of >20 units alcohol, diagnosis of a chronic medical condition (e.g. diabetes, cardiovascular or liver disease), or on a weight-loss program / not weight-stable for 3 months prior. Ethical approval was obtained from the East of England - Cambridge Central Research Ethics Committee and studies were conducted in accordance with the Declaration of Helsinki. All participants provided written informed consent.

### Protocol

The study design included 2 visits to the NIHR Cambridge Clinical Research Facility and Wolfson Brain Imaging Centre, 5 days apart. For Visit 1 (Day 0 13:00 to Day 1 09:00), participants arrived having had lunch and underwent two MR scans: one using a Siemens 3T Prisma (Erlangen, Germany) to perform proton magnetic resonance spectroscopy (¹H-MRS) to assess lipid content and composition, one using a Siemens 7T Terra (Erlangen, Germany) to perform deuterium magnetic resonance spectroscopy (²H-MRS) to assess baseline ^2^H in liver fat and water. In the Cambridge Clinical Research Centre (CCRC), following a brief rest, blood pressure and anthropometric measurements were taken. Baseline blood samples were then collected at 18:00, participants having fasted since lunch at around 12:00. Participants ate an energy-balanced evening meal around 18:30, then at 20:00 and 22:00 drank two priming doses of 70 % microbiologically-tested and filtered deuterated water, designed to achieve body water enrichment of 0.3 %. From this point until they returned on Day 5, participants were asked to make drinks substituting maintenance water (at 0.45 % enrichment) for normal water, in order to maintain stable body water enrichment. They were provided with a daily diary to document the number of maintenance water bottles consumed and whether they had consumed any liquids not made from maintenance water. At 08:00 the following morning (12 h after initial deuterated water priming), fasting blood samples were taken prior to breakfast before participants left. They returned on Day 5 in the afternoon; a blood sample was collected and another 7T ²H-MRS scan performed to enable assessment of the stored liver lipids formed by DNL.

### MRS and MRI Acquisition

#### ²H-MRS Liver

A custom-designed coil was used: a rigid ^2^H/^1^H transmit-receive surface coil (RAPID Biomedical, Germany) with ^2^H in quadrature (overall 23cm by 11cm) and ^1^H as a single elliptical loop of 17cm by 11cm. The 11cm dimension of the coil was designed to maximise signal from the liver and minimise that of visceral adipose tissue. The coil was superior within the housing such that coil positioning to the inframammary fold on females would permit optimal sensitivity of liver. The optimal curvature of the coil design was determined using previous imaging of people with varying BMI, with the coil was placed on the right anterior abdomen to maximise the sensitivity region to the liver. Three phantoms within the coil housing aided consistent positioning between visits. One of these was weakly dosed with deuterated water and used as an external standard to enable comparison between visits; its cylindrical shape caused a geometric susceptibility efect that moved its resonance to 7.4 ppm. Chemical shift imaging (CSI-fid) with weighted k-space acquisition was used to acquire spatially localized spectra (field of view (FOV) 260x280x240 mm, 13x14x12 scan resolution, Hamming-filtered and interpolated to 16x16x16) covering the right anterior abdomen. Voltages were calibrated and chosen such that the overall water signal intensity from a fid sequence (repetition time (TR) 900ms) was just past the maximum, so that signal intensity would be optimised approximately 1 voxel into the liver, away from subcutaneous fat and muscle. Using this technique no signal bleed from subcutaneous or visceral fat was detected in the liver. A preliminary inversion recovery test suggested the T1 relaxation time of ^2^H liver water was ∼270 ms (data not shown), and so TR 700ms was chosen for determination of the water signal intensity. In total there were four CSI-fid datasets, one with TR 700ms and three with TR 330ms, with scan times 13.52, 6.37, 12.88 and 25.97 mins respectively. All four were combined in such a way to optimise signal to noise and the resultant spectra used for determination of fat signal intensity. On the Day 5 return visit the coil and CSI grid were carefully repositioned exactly as for the baseline scan, using distance measurements to sternal notch and anatomical references relative to the 3 coil phantoms.

### 1H-MRS Liver

T2-weighted HASTE images were acquired in exhale to enable positioning a single 30×30×30 mm voxel in a similar location to the ^2^H acquisition, avoiding gall bladder and biliary tree, and adjacent adipose tissue, lung, and bowel. B₀ shimming was optimized using the GRE abdomen routine. Spectra were acquired using the STEAM sequence (TR 7 s, echo time (TE) 20 ms), with water suppression and 65 signal averages, participants performing shallow breathing following standardized instructions, so data was acquired in expiration. A non-water suppressed spectrum (4 averages) using the same acquisition parameters was used to assess water. To assess transverse relaxation times (T₂), non–water-suppressed spectra (4 averages) were acquired with TE 20, 50, 70, 90, 130, and 180 ms.

### 1H-MRS Soleus Intramyocellular Lipid

A T1-weighted turbo spin-echo sequence was used on the right calf to identify the optimal slice and voxel location within the soleus muscle. Water-suppressed and non–water-suppressed spectra were obtained from a voxel of cube length 13 mm, using the PRESS sequence with TE 35 ms and TR 5 s. Signal was acquired with 32 averages for the water-suppressed scan and 4 averages for the non–water-suppressed scan.

### 1H-MRI Visceral and Subcutaneous Adipose Tissue

Abdominal subcutaneous (SCAT) and visceral adipose tissue (VAT) volumes were assessed using the DIXON VIBE (Volumetric Interpolated Breath-hold Examination) MRI sequence. A slab of 18 axial slices centred on the L4 vertebral level was acquired (TE₁/TE_2_ 1.23/2.46 ms) with no fat or water suppression, and the manufacturer’s generated fat image used for calculation of VAT and SCAT area. Acquisition parameters were: single average, 500 mm FOV, 3.0 mm slice thickness, 20 % gap, and base resolution 320. Regional MRI analysis of the slice centred at the L4 vertebral level was performed using Analyze software (AnalyzeDirect, Overland Park, KS).

### 1H-MRS Visceral and Subcutaneous Adipose Tissue

A non-water suppressed spectrum from visceral and subcutaneous adipose tissues was acquired using STEAM (TR 7 s, TE 20 ms), with 12 and 8 averages and sample volumes ∼0.5 and ∼1 cm^3^, respectively. To assess the T₂ relaxation time of subcutaneous adipose tissue, taken as an indirect increasing measure of subcutaneous adipocyte size (19), single-average spectra were acquired at additional TE of 50, 70, 90, 130, and 180 ms, using the same localization and acquisition parameters.

### MRS Spectral Processing and Quantification

#### 2H-MRS Liver

CSI spectral data were opened using the Open-Source eXtensible Spectroscopy Analysis (OXSA) MATLAB code (33) and superimposed onto the corresponding anatomical image. The TE700 ms dataset was loaded and the integration function in MATLAB was used to generate water signal intensity heatmaps. These were used to identify the slices and voxels for signal extraction (typically 42-81 voxels), spectra from which were frequency-aligned and averaged in jMRUI v6.0 (34, 35). The same procedure was applied to the other 3 CSI datasets (with TE 330 ms), resulting in 3 additional spectra. The 4 spectra were then averaged in a specific order to generate a final consolidated spectrum optimised for signal-to-noise. The same post-processing workflow was applied to the post-dosing dataset using the identical voxel positions as in the pre-dose scan. The external phantom was used to normalise signals between visits.

The consolidated and TR 700ms spectra had 1.5 Hz line broadening applied and were phase-corrected prior to being baseline-corrected (36) in MATLAB using a spline function defined by multiple points throughout the spectrum known to be part of the baseline. Frequency ranges corresponding to the water (TR 700ms spectra) and fat peaks (consolidated spectrum) were identified, and integration used to quantify signal intensities. The same frequency ranges were used for the post-dosing dataset.

Precision of the ^2^H fat measures was assessed by comparison of ^2^H fat from the 25.97 min scan (termed ‘scan 1’) with the optimally-combined other 3 scans (‘scan 2’).

To derive MR-specific equations for assessing liver lipid from DNL, we start by using an equation for fractional DNL from Delgado et al (37):

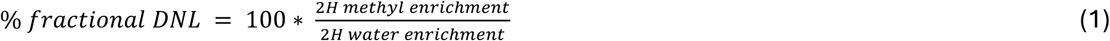

And, importantly, we define enrichment as:

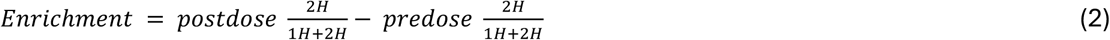

If we assume the total (^1^H + ^2^H) lipid and water pool does not alter over the 5 days, and is approximated to the pre-dose ^1^H signal, and ^2^H signals are normalised to the external phantom (vial), then:

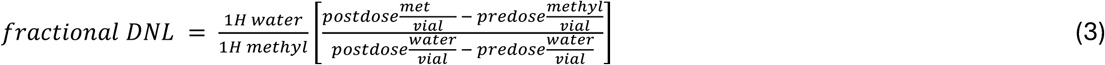

However, as we cannot distinguish methyl from methylene signals, equation (3) is adapted for combined methyl + methylene signals. In addition, a factor is introduced to account for the maximum incorporation number (N) of deuterium atoms in palmitate. In vivo, N has been shown to be 22 in methyl esters of palmitate (28, 38), yielding a maximum enrichment from de novo synthesis of 65 % (28). We therefore derive:

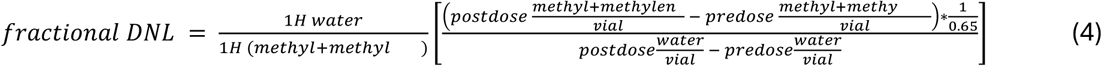

Then converting to an absolute amount of lipid formed by DNL, termed Absolute DNL, in units of liver % weight/weight,

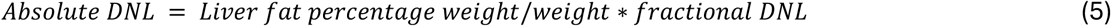

where liver fat percentage weight/weight is the ^1^H lipid methylene relative to the sum of methylene and water (39).

### 1H-MRS Liver, Visceral and Subcutaneous Adipose Tissue

^1^H spectra from the liver, SCAT and VAT were processed in jMRUI v 6.0 (34, 35). Individual averages were frequency-aligned before being averaged. Spectral fitting was conducted using the AMARES algorithm (40) with identical prior-knowledge parameters across datasets. In liver water-suppressed and SCAT & VAT spectra, the water peak was excluded from fitting and 6 key resonances were modelled between 0 and 3 ppm: methyl (∼0.9 ppm), methylene (∼1.3 ppm), γ-methylene (∼1.6 ppm), allylic (∼2.02 ppm), α-carboxyl peak (∼2.2 ppm), and diallylic (∼2.74 ppm). All peaks had soft constraints on their frequencies and a fixed relative phase of zero. The α-carbonyl amplitude was fixed relative to the methyl, similar to (12) but using the theoretical 0.67. The allylic peak linewidth was constrained to 1.32 times that of the methyl peak. All resonances were modelled using Gaussian line shapes; for liver non–water-suppressed spectra, an additional Lorentzian peak at 4.7 ppm representing water was fitted without any prior-knowledge constraints.

Lipid composition was calculated based on Roumans et al (12):

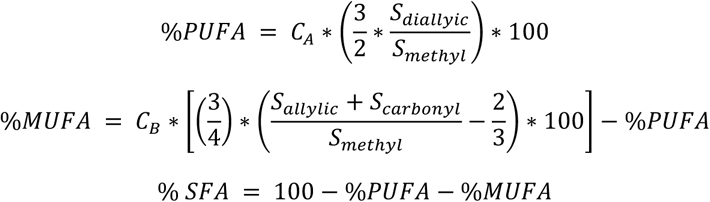

where S is the signal for each resonance, and CA and CB are 0.83 and 0.99, respectively, for TE20ms using STEAM at 3T (12).

Absolute quantification of liver fat (termed IHL herein) was expressed as percentage weight/weight, [IHL] = 100 * (So methylene) / (So methylene + So water) as demonstrated within Buitanga et al. (39), where So is the T_2_-corrected signal intensity of the resonance. T₂ relaxation times for liver water, CH₂ and CH₃ signals and SCAT CH_2_ were quantified from the slope of the plot of natural log-transformed signal intensity versus TE over the range of TE from 20 to 180 ms. There were no statistical between-individual differences in liver T_2_, so average values (from 5 SA and 4 E) of T_2_ = 32, 57, 50 ms for water, fat methylene, and fat methyl, respectively, were used for T_2_ correction.

In order to compare the amounts of liver fat derived from ^1^H and natural abundance ^2^H, liver fat was also expressed as (S_o methyl_ + S_o methylene_)/S_o water_ for both ^1^H and ^2^H (as ^2^H fat signal contains both methyl and methylene protons), where So is the T_2_-corrected signal intensity of the resonance (^1^H) without proton density correction.

### 1H-MRS Soleus Intramyocellular Lipid

Spectra were analysed in jMRUI v6.0 (34, 35) using the AMARES (40) algorithm with prior knowledge parameters as outlined previously (41). In short, soft constraints were applied to the frequencies and linewidths of CH₂ peaks, while CH₃ peak parameters were constrained relative to CH₂ resonances, in accordance with established prior knowledge. Both CH₂ and CH₃ signals were quantified relative to the unsuppressed water peak with all amplitudes estimated during fitting.

### Plasma DNL measures

The incorporation of deuterium into plasma TG during ingestion of deuterated water was used to determine the fractional DNL_plasma_ as previously described (42). Total plasma lipids were extracted using chloroform-methanol, and TAG was separated by solid phase extraction. Fatty acid methyl esters were prepared using methanolic sulphuric acid, and fatty acid relative abundance (mol%) was determined by gas chromatography. DNL was assessed based on the incorporation of ^2^H in plasma water into plasma TAG palmitate using GC-MS, monitoring ions with mass-to-charge ratios of 270 (Mþ0) and 271 (Mþ1). % fractional DNL was then defined as 100 x plasma palmitate enrichment/(22 × plasma deuterium enrichment) (43). Enrichment was defined as the enrichment above baseline levels, and Day 5 water enrichments assumed a linear change in ^2^H water enrichment between Day 1 and 5.

### Biochemical assays

Fasting biochemical measures were measured by standard clinical laboratory assay methods as follows. Triglyceride, cholesterol (total and HDL), GGT, ALT, AST and glucose were analysed using the Siemens Healthineers Dimension EXL analyser and reagents. Cholesterol (LDL) was then calculated using the Friedewald equation. Insulin was analysed using the DiaSorin Liaison XL analyser and reagent. Adiponectin and leptin were analysed using in-house immunoassays based on the Revvity DELFIA platform utilising Bio-Techne R&D Systems antibodies and standards. B-hydroxybutyrate was analysed using Stanbio reagents; NEFA using Roche reagents. HbA1c was analysed using the Tosoh G11 analyser.

The homeostatic model assessment for insulin resistance (HOMA-IR) was calculated as the product of fasting insulin (mU/L) and fasting glucose (mmol/L), divided by 22.5. Insulin was divided by 6.0 to convert pmol/L to mU/L.

### Statistical methods

Statistics were performed in SPSS v28. Mann U Whitney test was used for between-group comparisons, absolute intraclass correlation coefficient (ICC) for ^2^H fat reproducibility, and Spearman’s correlation coefficient for associations with other variables.

## Data Availability

Datasets from participants who consented to data sharing are available from the corresponding author upon reasonable request.

## Acknowledgements

We thank the study participants, staff at the Wolfson Brain Imaging Centre and the NIHR Cambridge Clinical Research Facility. We are grateful to everyone at RAPID Biomedical GmbH (Rimpar, Germany) for custom design of the excellent ^2^H/^1^H coil. We would like to acknowledge staff at the NIHR Core Biochemistry Assay Laboratory, Cambridge Biomedical Research Centre for performing all the assays other than the HbA1c, which we acknowledge the support and analytical services of Cambridge University Hospitals NHS Foundation Trust Laboratory Research Services and clinical laboratories.

This work was funded by the Medical Research Council [MR/V011758/1 to AS], and the study used infrastructure supported by the NIHR Cambridge Biomedical Centre [grant numbers BRC 1215 20014, NIHR203312]; the Wellcome Trust [grant numbers 208363/Z/17/Z, 226800/Z/22/Z]; and the Medical Research Council [grant numbers MC_UU_00014, MC_UU_00039]. The GC-MS analysis was funded by the British Heart Foundation [Fellowship FS/SBSRF/21/31013 to LH]. KMB is currently in receipt of a Hans Fischer Senior Fellowship at the Technical University Munich. NGF acknowledges funding support by MRC Epidemiology Unit [MC_UU_00006/3], the NIHR Cambridge Biomedical Research Centre, and NIHR Senior Investigator Award [NIHR202397]. DBS is supported by the Wellcome Trust [WT 219417], the MRC [MR/X00970X/1], and NIHR Cambridge Biomedical Research Centre and NIHR Rare Disease Translational Research Collaboration. AS gratefully acknowledges support from the NIHR Cambridge Clinical Research Facility. This is a summary of independent research funded by the Medical Research Council and carried out at the National Institute for Health and Care Research (NIHR) Cambridge Clinical Research Facility (CRF). The views expressed are those of the author(s) and not necessarily those of the Medical Research Council, the NIHR or the Department of Health and Social Care.

