## Supplementary S1 for "Non-invasive ^2^H-MRS reveals a greater liver fat contribution from de novo lipogenesis in South Asians compared with Europeans"

**Supplementary Information**

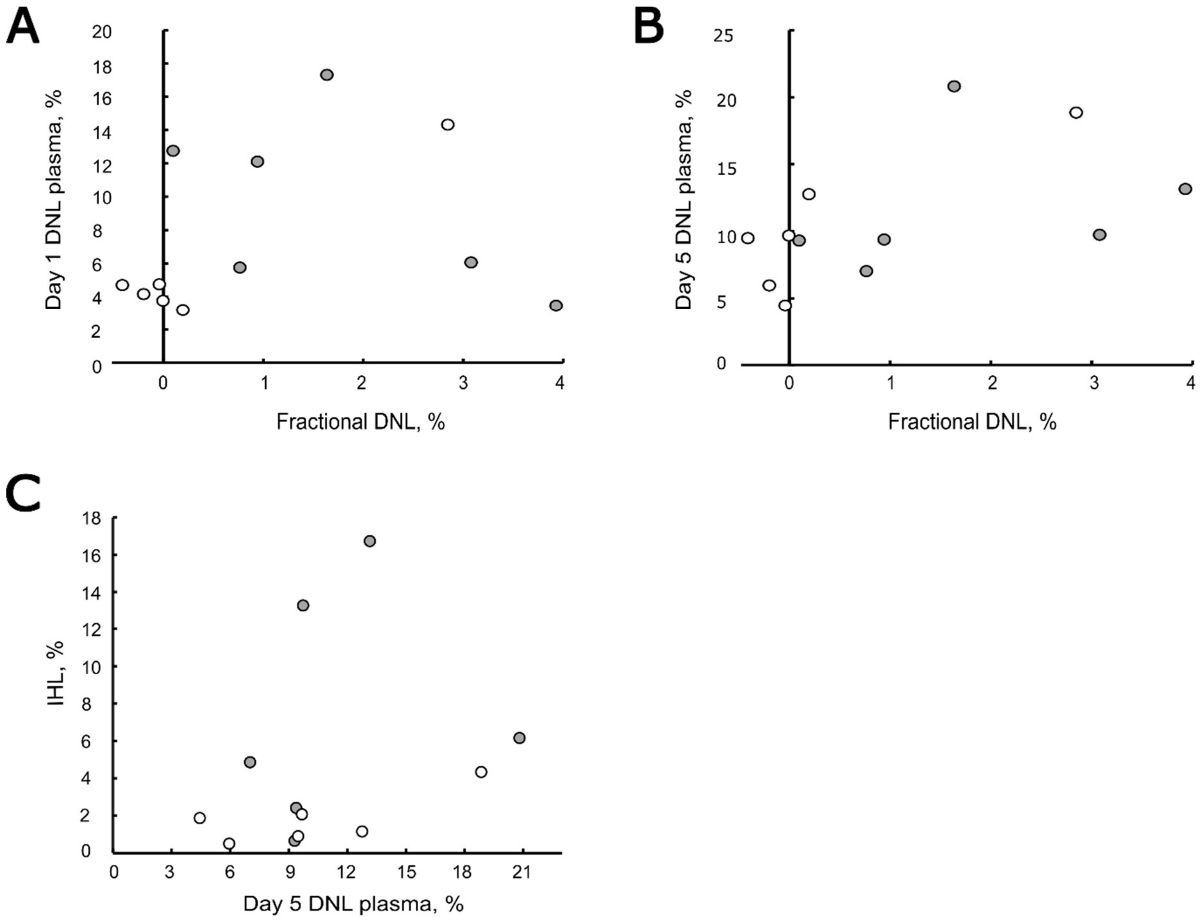


**Figure S1. Comparison of traditional DNL measures (DNL plasma) with the new MR measure of fractional liver fat originating from DNL (fractional DNL).** 'Traditional' DNL($DNL_{plasma}$) assesses fractional $^{2}$H-palmitate within plasma triglyceride. $DNL_{plasma}$ assessed on Day 1, 12 hours after priming dose (A), and Day 5 (B), vs MR-measured fractional liver fat originating from DNL on Day 5. (C) Liver fat content (IHL) vs Day 5 $DNL_{plasma}$. Data are for South Asian (SA, grey circles) and Europeans (E, white circles).
